# Laboratory based surveillance of *SARS-CoV-2* in Pakistan

**DOI:** 10.1101/2020.06.10.20126847

**Authors:** Nazish Badar, Aamer Ikram, Hamza Ahmad Mirza, Abdul Ahad, Muhammad Masroor Alam, Yasir Arshad, Massab Umair, Salmaan Sharif, Afreenish Hassan, Muhammad Salman

## Abstract

COVID-19 cases are alarmingly increasing in Pakistan since May 2020. Laboratory based surveillance system has been in place since the start of the pandemic. The genomic surveillance of SARS-CoV-2 strains isolated locally has been conducted based on partial ORF1b. The sequences were classified to show the phylogenetic correlation and showed 100% homology with those detected in neighboring countries India and China. The rapid increase in cases has led to development of robust strategies to enhance the laboratory testing capacity. We are currently meeting the country requirement to diagnose the virus in the community. Nonetheless, factors like recent ease in lockdown measures has led to massive rise in number of cases in few weeks time.

## Manuscript

The world is currently going through one of the major public health challenges of twenty first century: *SARS-CoV-2* pandemic that emerged from Wuhan, China during December 2019. Pakistan has joined the growing number of Asian countries reporting cases of novel coronavirus disease (COVID-19), starting with two confirmed patients on 25^th^ Feb 2020 and since then, the situation is propagating atrociously. As of May 21 2020, 50694 laboratory confirmed cases and 1067 deaths (2.14% case fatality rate) have been reported in Pakistan (Figure 1B and 1C).

**Figure 1.**
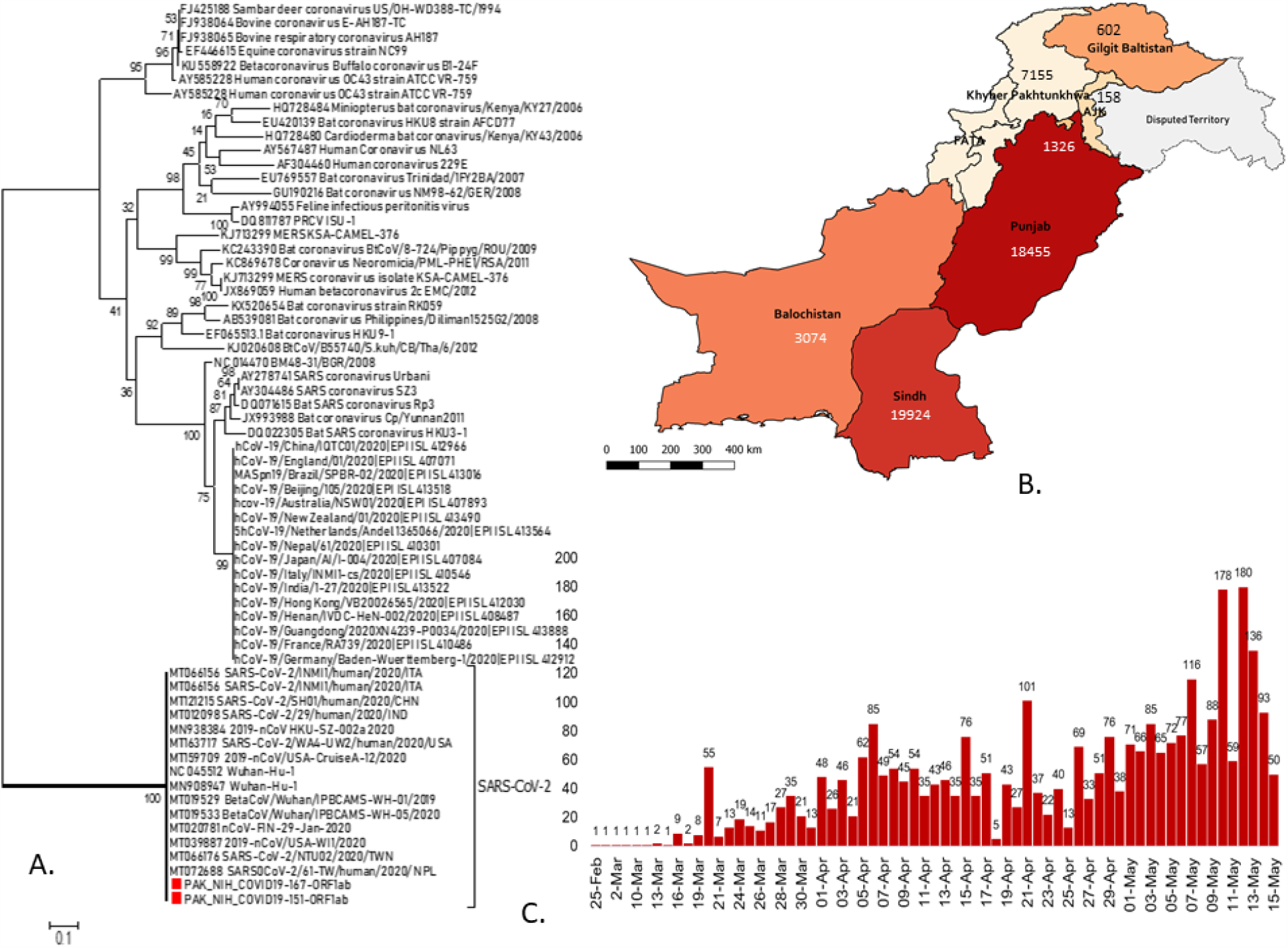
A. Phylogenetic tree of SARS-CoV-2 strains based on ORF1ab detected in Pakistan (red box indciates first two cases identified in Pakistan) B. Confirmed case count mapped across provinces in Pakistan as of May 21 2020 C. Number of confirmed SARS-CoV-2 samples tested at National Institute of Health, Islamabad, Pakistan as of may 21 2020.

Since February 2020, Pakistan has initiated hospital-based surveillance system for SARS-CoV-2 infections, mainly targeting tertiary care and teaching hospitals across Pakistan. These hospitals serve as the filter clinic for initial screening of patients based on their clinical signs and symptoms and act as STAT labs to collect and refer specimen to designated laboratories. The diagnostic capacity in Pakistan has been timely strengthened by engaging laboratories from both public and private sector and currently expanded to 70 facilities. The vertical program already functional in the country including (but not limited to) polio, measles, rotavirus, influenza and tuberculosis supported the government by transforming their infrastructure and allocating human resource for COVID-19 laboratory diagnostics services.

In Pakistan, the National Institute of Health, Islamabad (NIH) is the only national reference public health laboratory in the country coordinating a lab-based systematic influenza surveillance network at seven sentinel sites supported by the Centers for Disease Control and Prevention, USA. The NIH, Pakistan initiated the SARS-CoV-2 confirmation using real-time PCR assay following protocol recommended by the World Health Organization targeting E gene of SARS-CoV-2 strain (1). The NIH based COVID-19 laboratory has extensive diagnostic capacity and tested 36,500 clinical samples with detection in 3019 (8.27%) confirmed cases. Out of 36,500 specimens, 68% (24820) individuals were reported without history of any symptoms and identified through contact tracing or epidemiological investigations. Only 21% cases were presented with influenza like illness (fever, cough, sore throat) and 11% with severe acute respiratory infection that fulfilled COVID-19 cases definition [2]. These findings are suggestive of superspreading phenomenon in which few individuals unusually infect large number of secondary cases. These facts also highlight the significance of both laboratory and community-based surveillance to reveal the extent of transmission in our settings where symptomatic patients are under-reported or missed by the national surveillance system.

In addition to diagnostic services, the NIH Pakistan is supporting the national authorities to monitor the extent of community transmission in the country by genomic surveillance of SARS-CoV-2 strains and their associated epidemiological information. The local strains based on partial ORF1b sequence were classified into A3 clade and showed 100% homology with those detected in neighboring countries India and China (Figure 1A). The very first cases in the country had history of travel to Iran where thousands of pilgrims visit every year performing their religious activities. The initial visitors then returned to Pakistan at the time when screening and quarantine measures were not fully adopted by Pakistan. Likewise, the spurred rise in COVID-19 cases in Pakistan are linked to a mass religious gathering in Lahore held during 11-12 March 2020 attended by at least 150,000 nationals as well as international clerics. The infected attendees then moved across the country for preaching missions thus igniting the cryptic virus transmission until the symptomatic cases started appearing every so often (3). The current transmission trends across the country seems reasonable as revealed by the epidemiological tracking and expansion of cases seeded by the local religious congregations and social gatherings and later by relaxing the lock-down situation.

## Data Availability

Not applicable

## References

1. Corman VM, Landt O, Kaiser M, et al. Detection of 2019 novel coronavirus (2019-nCoV) by real-time RT-PCR. Euro Surveill. 2020;25(3):2000045. doi:10.2807/1560-917.ES.2020.25.3.2000045.

2. World Health Organization. Coronavirus disease (COVID-19) technical guidance: Surveillance and case definitions. Available at: https://www.who.int/emergencies/diseases/novel-coronavirus-2019/technical-guidance/surveillance-and-case-definitions. Access date: 21 May 2020

3. Nishimura et al. Closed environments facilitate secondary transmission of coronavirus disease 2019 (COVID-19). medRxiv preprint doi:https://doi.org/10.1101/2020.02.28.20029272.

